# The immediate and longer-term impact of the COVID-19 pandemic on the mental health and wellbeing of older adults in England

**DOI:** 10.1101/2021.04.30.21256385

**Authors:** P Zaninotto, E Iob, P Demakakos, A Steptoe

## Abstract

**Objective:** To evaluate changes in mental health and wellbeing before and during the initial and later phases of the COVID-19 pandemic and investigate whether patterns varied with age, sex, and socioeconomic status.

**Design:** Prospective cohort study.

**Participants:** English Longitudinal Study of Ageing cohort of 5146 community dwelling adults aged 52 years and older (53% women, average age 66.74 years, standard deviation 10.62) who provided data before the pandemic (2018-19) and at two occasions in 2020 (June-July and November-December).

**Main outcome measure:** Depression, poor quality of life, loneliness and anxiety.

**Results:** The prevalence of clinically significant depressive symptoms increased from 12.5% pre-pandemic to 22.6% in June-July 2020, with a further rise to 28.5% in November-December. This was accompanied by increased loneliness and deterioration in quality of life. The prevalence of anxiety rose from 9.4% to 10.9% between June-July and November-December 2022. Women and non-partnered people experienced worse changes in mental health and wellbeing. Participants with less wealth had lowest levels of mental health before and during the pandemic. Higher socioeconomic groups had better mental health overall, but responded to the pandemic with more negative changes. Patterns of changes were similar across age groups, the only exception was for depression which showed a smaller increase in the 75+ age group than in the youngest age group (50-59 years).

**Conclusions:** These data showed that mental health and wellbeing continued to worsen as lockdown continued, and that socioeconomic inequalities persisted. Women and non-partnered people experienced greater deterioration in all mental health outcomes. The immediate provision of diagnosis of mental health problems and targeted psychological interventions should target and support sociodemographic groups of older people at higher risk of psychological distress.

**What is already known on this topic:**

- The COVID-19 pandemic and mitigation measures have upended the economic and social lives of many, leading to widespread psychological distress.
- During the early months of the pandemic, levels of depression, anxiety and loneliness were high and lower levels of wellbeing were reported across the adult population, with certain higher risk groups identified.
- However, evidence from longitudinal studies of representative samples of older adults that include pre-pandemic data is scarce, and little is known about mental health beyond the initial period of the pandemic. Repeated assessments are needed in order to understand whether mental health and wellbeing levels recovered or continued to deteriorate throughout 2020.

**What this study adds:**

- These data suggest that mental health and wellbeing deteriorated significantly during June-July 2020 compared with pre-pandemic levels and continued to deteriorate during the second national lockdown in November-December 2020, showing that older individuals did not adapt to circumstances.
- Inequalities in experiences of mental ill-health and poor wellbeing during 2020 were evident, with women, individuals living alone and those with less wealth being particularly vulnerable. Furthermore, socioeconomic inequalities in mental health have persisted during the pandemic.

## Introduction

It is now well documented that the COVID-19 pandemic has significantly affected the mental health and wellbeing of the adult population, not only in the UK and other high income countries, but in low and middle oncome countries as well. ^1 2^ Stress directly related to the disease and worries about disruption to healthcare services, employment, financial security and limitations to social contacts are likely to contribute to psychological distress. Much of the evidence has derived from internet studies initiated after the onset of the COVID-19 lockdown. Young adults, women, people of lower socioeconomic status (SES), ethnic minorities, and individuals with pre-existing mental and physical health problems appear especially vulnerable. ^3-6^ Such studies lack information about mental health and wellbeing before the pandemic, so increases in distress are inferred rather than measured directly. Data collection online rules out participants who do not have internet access, including socially marginalised groups and sectors of the older population. ^7^ Nevertheless, these findings have been corroborated by longitudinal studies that compare experiences during the early months of the pandemic with data collected in past years. ^8^ Analyses of the UK Longitudinal Household Study (UKHLS) confirm increases in distress measured with the General Health Questionnaire (GHQ) during the early months of the pandemic and lockdown in the UK. ^8 9^, consistent with studies in other countries. ^10 11^

Older adults are at increased risk of serious illness and death following COVID-19 infection ^12^, and are particularly vulnerable to social isolation and loss of access to social and health care. These experiences may lead to poor mental health and wellbeing that are in turn associated with cognitive decline ^13^ incident dementia ^14^ mortality ^15 16^ and several physical health conditions. ^17 18^ Despite these factors, longitudinal analyses indicate that the impact on mental health has been smaller among older than younger adults. ^19^ Indeed, some studies have reported there is little increase in depression or deterioration in wellbeing among older people. ^8 20 21^ However, most studies have been conducted in the early months of the pandemic and associated stay at home directives, and these patterns may change with repeated social distancing regulations as have occurred in the UK in the past year. Prolonged restrictions and persistence in infection rates may have taxed older people’s capacity to adapt, resulting in increasing levels of mental distress.

Assessments of mental health and wellbeing in the English Longitudinal Study of Ageing (ELSA) carried out in June-July and November-December 2020 permitted analysis of the impact of the COVID-19 pandemic in a large representative sample of men and women aged 52 and older. Data were collected both on line and by telephone interview, so that participants who were unable to access the internet were not excluded. We evaluated whether mental health and wellbeing were affected at two time points during the pandemic compared with previous years, using standardised measures of depressive symptoms and anxiety to evaluate mental health, with assessments of quality of life and loneliness as measures of wellbeing. We also investigated whether patterns varied with age, sex, and SES, as found in surveys that have involved adults throughout the age spectrum.

## METHODS

### Sample

The data came from the COVID-19 Substudy of the English Longitudinal Study of Ageing (ELSA). ELSA is an ongoing prospective population-based cohort study of older adults aged 50 years and over living in England ^22^. The study began in 2002 with participants drawn from the Health Survey for England (HSE). This sample was representative of the general English population of older adults ^23^. Data on several health and socioeconomic outcomes have been collected every two years, with additional nurse visits every four years for the collection of biomedical data. There have been nine waves of data collection since the study began in 2002/03 (i.e. wave 1). The ELSA COVID-19 Substudy is a follow-up study based on the regular ELSA sample. It has been launched in 2020 to investigate the socioeconomic and psychological impact of the COVID-19 pandemic on the 50+ population of England. The first wave of data collection took place in June-July 2020, and the second wave was completed in November-December 2020 ^24^. The response rate was high in both waves (75%). The longitudinal response rate was 94.2%. For the purpose of the present study, we created a longitudinal sample including 5,146 core ELSA members of the COVID-19 Substudy who participated in both COVID-19 waves and in a regular ELSA wave previous to COVID-19 (either wave 9 (2018/19) or 8 (2016/17)). All analyses were weighted to match the latest population estimates for age, sex, and region in England and account for non-response to the ELSA COVID-19 Substudy survey ^24^. All respondents provided informed consent. Further information regarding the sample design and data collection methods can be found on the study website (https://www.elsa-project.ac.uk/).

### Measures

#### Outcomes

We focused on the following mental health outcomes: depression, quality of life, loneliness, and anxiety. Depressive symptoms were ascertained using the 8-item Centre for Epidemiological Studies Depression (CESD-8) scale, which measures eight different symptoms of depression (e.g. “felt depressed”, “everything I did was an effort”, “sleep was restless”). This scale has previously been validated against gold-standard psychiatric interviews with good sensitivity and specificity ^25^. A dichotomous (yes/no) response was used for each item, resulting in a total CESD-8 score ranging between zero (no symptoms) and eight (all eight symptoms). We then created a binary variable using a cut-off point of four or more symptoms to identify likely cases of clinical depression, which is equivalent to the conventional threshold of 16 or higher on the full 20-item CESD scale ^26^. Quality of life was measured using the 12-item version of the Control, Autonomy, Self-realisation, and Pleasure (CASP) scale, a self-completion questionnaire that has been developed to assess the quality of life and wellbeing of older people. The 12-item version of CASP measures three domains of quality of life, including ‘Control and Autonomy’ (e.g. “My age prevents me from doing the things I would like to do”), ‘Pleasure’ (e.g. “I look forward to each day”), and ‘Self-realisation’ (e.g. “I feel that life is full of opportunities”) ^27^. Each item is scored on a 4-point scale (“Often”, “Sometimes”, “Not often”, “Never”). The resulting item scores were summed to create an index of quality of life where higher scores indicate poorer wellbeing (range: 1-48). Loneliness was assessed using the 3-item revised University of California (UCLA) Loneliness scale ^28^ (“How often do you feel”: 1) “lack of companionship?”; 2) “left out?”; 3) “isolated from others?”), and an additional item asking participants how often they feel lonely. Each question was rated on a 3-point scale (1 = “hardly ever/never”; 2 = “some of the time”; 3 = “often”). The individual item scores were then summed together to produce a total score, with higher values indicating greater loneliness (range: 1-12). Anxiety was measured using the 7-item generalised anxiety disorder scale (GAD-7), which evaluates the presence of various symptoms of generalised anxiety disorder (GAD) (e.g. “Feeling nervous, anxious or on edge”, “Not being able to stop or control worrying”). This scale has been shown to be valid and reliable tool for screening for GAD and to assess its severity in both research and clinical practice.^29^ Each item is scored on a 4-point scale (“Not at all, “Several days”, “More than half the days”, “Nearly every day”). We used a total score of 10 or greater as a cut-off point for identifying cases of GAD. To understand the impact of the COVID-19 pandemic on depression, quality of life, and loneliness, we compared the participant’s scores at the two COVID-19 waves with those of their most recent assessment before the start of the pandemic (i.e. wave 9 or 8). For anxiety, we only compared the change between the two COVID-19 waves, as this scale was not included previously in the regular ELSA survey.

#### Sociodemographic characteristics

The main sociodemographic characteristics considered in the analysis were: age, sex, wealth, and marital/partnership status. The variable age included three groups: 50-59, 60-74, and 75+ years. Sex was a binary variable (men/women). Wealth was derived from a comprehensive assessment of the participant’s economic resources (e.g. financial, housing, and physical wealth) excluding pension wealth, and was categorised into tertiles (1^st^ = lowest wealth; 3^rd^ = highest wealth). Partnership status was a binary variable indicating whether the participant had a partner. We also presented descriptive statistics for the following characteristics: ethnicity (white/other), education (“low” = Compulsory School Leaving/ “medium” = A-levels & College/ “high” = Degree or above), employment status (employed/ retired/ other not working), home tenure (owns outright/ owns with mortgage/ rents), and limiting longstanding illness (no/ yes). Age and partnership status were measured at the first COVID-19 assessment, while wealth, education, employment status and limiting longstanding illness were determined in pre-pandemic assessments (i.e. wave 9 or 8). Further, we derived a binary variable indicating whether the participant had experienced COVID-19 at the first or second COVID-19 wave. The following criteria were applied to identify confirmed or suspected cases of COVID-19: participants were found to be COVID-19 positive on testing, or were hospitalised due to COVID-19, or reported two of the three core symptoms as defined by the UK National Health Service (NHS) (i.e. high temperature, a new continuous cough, and loss of sense of smell or taste).

### Statistical analyses

We examined the effect of the COVID-19 pandemic on changes within an individual’s mental health using two-way fixed-effects regression models. This modelling approach can control for all unobserved confounders that vary across individuals but are constant over time (e.g. genetic susceptibility), and those that are constant across individuals but change over time ^30^ (e.g. time-period effects). Hence, it can provide more reliable estimates of the plausible causal effect of the COVID-19 pandemic on changes in mental health. A linear model was used to estimate changes in the total scores of quality of life and loneliness, as well as in the probability of the binary depression and anxiety scores. For the binary outcomes, a linear probability model was chosen over a logistic fixed-effects model since this approach would exclude those who had concordant scores across all waves thereby affecting the representativeness and statistical power of the analysis. Firstly, we estimated the effect of the COVID-19 pandemic on depression, quality of life, and loneliness using two binary independent variables indicating whether the outcome was measured at the first or second COVID-19 wave (vs before COVID-19). For anxiety, we used one binary variable indicating whether the outcome was measured at the second COVID-19 wave (vs first COVID-19 wave). We then repeated this analysis using the standardised outcome scores to enable direct comparisons across the outcomes. Secondly, we tested interaction effects between a COVID-19 period indicator for whether the outcome was measured before or during COVID-19 (wave 1 or 2) and the four sociodemographic factors described above to understand whether/ how the average change in each mental health outcome before and during the COVID-19 pandemic might vary across different sociodemographic groups. For each outcome, we first tested each interaction effect individually, and then fitted a mutually adjusted model including all interactions between the COVID-19 period indicator and the sociodemographic factors. For the significant interaction effects found in the mutually adjusted models, we then produced graphs of the predicted outcome values by the selected sociodemographic groups. The percentage of missing data in the variables ranged between 0 and 6%. In addition, due to a survey error, for around 75% of the sample the last item of the CESD-8 questionnaire was not administered at the first COVID-19 wave. This type of missing data is classified as missing completely at random (MCAR), and can be dealt efficiently with multiple imputation (MI).^31^ We used MI by chained equations with all variables of the analysis included as predictors of the imputation models as well auxiliary variables. We created twenty imputed datasets, and then pooled the regression estimates across the imputed datasets using Rubin’s rules ^32^. The distribution of the variables in the imputed and observed data was similar, suggesting that the MI procedure produced accurate model estimates (see Table1 for sample characteristics). As sensitivity analysis, we restricted the analyses of the regression models on the sample of participants with complete data on all variables and compared these to the imputed results. Further, we repeated the main imputed analyses excluding participants who experienced COVID-19 at the first or second COVID-19 wave. Data management and regression analyses were conducted in Stata 16. Graphical and MI analyses were performed in R version 4.0.2, with the packages *mice* ^*33*^ and *ggplot2* ^*34*^.

**Table1.** Characteristics of the longitudinal ELSA COVID-19 sample (N=5146).

|  |  |
| --- | --- |
| **Sex** |  |
| Men | 47.10% |
| Women | 52.90% |
| **Age group** |  |
| 52-59 | 31.90% |
| 60-74 | 44.30% |
| 75 and over | 23.80% |
| **Ethnicity** |  |
| White | 92.80% |
| Other | 7.20% |
| **Partnership** |  |
| Partnered | 75.40% |
| Non partnered | 24.60% |
| **Education** |  |
| High | 20.50% |
| Medium | 46.60% |
| Low | 32.90% |
| **Employment status** |  |
| Employed | 39.00% |
| Retired | 51.40% |
| Other not working<sup>c</sup> | 9.60% |
| **Wealth (tertiles)** |  |
| 1st tertile (Poorest) | 43.20% |
| 2nd tertile | 29.20% |
| 3rd tertile (Richest) | 27.50% |
| **Home tenure** |  |
| Owens outright | 59.90% |
| Owens with mortgage | 21.90% |
| Rents | 18.20% |
| **Limiting Longstanding Illness** |  |
| No | 48.40% |
| Yes | 51.60% |
| **Confirmed or suspected COVID-19 cases <sup>a</sup>** |  |
| No | 94.60% |
| Yes | 5.40% |
| **Elevated depressive symptoms (CESD-8 ≥ 4) <sup>b</sup>** |  |
| No | 77.40% |
| Yes | 22.60% |
| **Poor Quality of Life (CASP-12) <sup>b d</sup>** |  |
| Mean (SD) | 22.53 (6.53) |
| Range | 12- 47. |
| **Loneliness <sup>b</sup>** |  |
| Mean (SD) | 5.65 (2.065) |
| Range | 2. - 12. |
| **Anxiety (Gad-7 ≥ 10) <sup>b</sup>** |  |
| No | 90.60% |
| Yes | 9.40% |
| ***Note.** Results based on 20 imputed datasets; percentages and mean are estimated using sampling weights; SD = standard deviation. <sup>a</sup> Prevalence of COVID-19 cases across waves 1 and 2. <sup>b</sup> Mental health scores at the first COVID-19 wave. <sup>c</sup> Permanently unable to work, looking after home and family, currently out of work. <sup>d</sup> Higher scores indicate poorer QoL* |  |

### Patient and public involvement

No patients were involved in setting the research question or the outcome measures, nor were they involved in developing plans for design or implementation of the study. No patients were asked to advise on interpretation or writing up of the results. We do plan to disseminate the results of the research to the relevant patient communities.

## RESULTS

### Descriptive analyses

#### (1) Sample characteristics

The characteristics of the analytical sample at the first COVID-19 wave are reported in Table1. The sample included 5,146 participants (52.9% women). Ages ranged from 52 to over 90 years (mean 66.74, SD 10.62). The majority of participants had a white ethnic background (92.8%), and most had a partner (75.4%). Only 4 participants who had a partner were living alone, while all participants without a partner were living alone. More than half of the sample was retired (51.4%), and 30% of the sample had low levels of education, and around 40% of participants were in the poorest wealth group. Approximately half of participants reported a limiting longstanding illness. Descriptive statistics of the mental health outcomes across the waves are reported in the Supporting Information (SI) Appendix (sTable1).

#### (2) Mental health outcomes before and during COVID-19

The predicted trajectories of the mental health outcomes before (i.e. wave 9 or 8) and during the COVID-19 pandemic derived from the fixed-effects models are illustrated in Figure1 and reported in the SI Appendix (sTable2 and sFigure1). Compared with pre-pandemic levels, all mental health outcomes deteriorated during the first COVID-19 wave (May-June 2020), and continued to worsen through to the second wave (Nov-Dec 2020). The probability of depression increased from 12.5% (95%CI 11.5, 13.4) pre-pandemic to 22.6% (95%CI 21.6, 23.6) in the first COVID-19 wave, and to 28.5% (95%CI 27.6, 29.5) in the second wave. This was accompanied by increases in the mean total scores of loneliness (pre-pandemic: 5.50 95%CI 5.45, 5.54; COVID-19 wave 1: 5.65 95%CI 5.61, 5.68; COVID-19 wave 2: 5.75 95%CI 5.71, 5.78) and poor quality of life (pre-pandemic: 21.6 95%CI 21.48, 21.72; COVID-19 wave 1: 22.5 95%CI 22.43, 22.63; COVID-19 wave 2: 23.1 95%CI 22.97, 23.16). Further, there was an increase in the levels of anxiety in the sample across the two COVID-19 waves from 9.4% (95%CI 8.8, 9.9) to 10.9% (95%CI 10.3, 11.5). The observed changes in mental health before and during the COVID-19 pandemic mirrored those of the fixed-effects analysis (see sTable2).

**Figure1.**
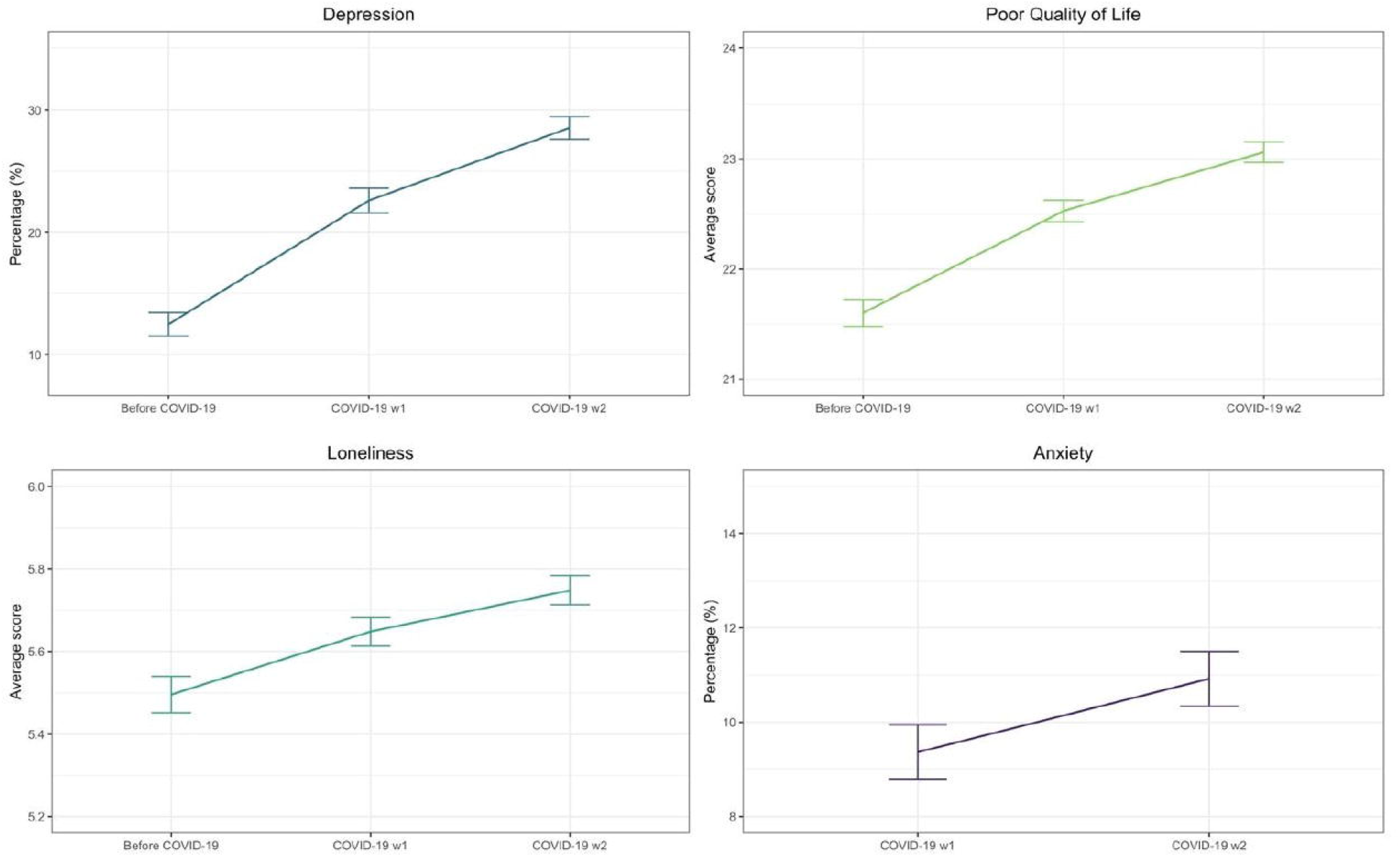
Predicted outcome trajectories before and during the COVID-19 pandemic (fixed-effects models). **Note**. ELSA COVID-19 longitudinal sample (N=5,146); weighted pooled estimates from two-way fixed-effects linear models across 20 imputed datasets.

#### (3) Impact of the COVID-19 pandemic on within-individual changes in mental health

Based on the fixed-effects models, we estimated the magnitude and statistical significance of within-individual changes in mental health before and during COVID-19. The estimated changes in mental health are reported in sTable3 (SI Appendix). During the first COVID-19 wave, the probability of depression increased by 10 percentage points (95%CI 0.09, 0.12), corresponding to an increase of 81% compared with pre-pandemic scores. Ratings of poor quality of life increased by 0.93 points (95%CI 0.73, 1.12), and loneliness increased by 0.l5 points (95%CI 0.08, 0.22), representing respectively an increase of 4.3% and 2.8% compared with pre-pandemic scores. Significant changes in mental health were also found in the second COVID-19 wave. Compared with the first COVID-19 wave, there was an increase of 6 percentage points (95%CI 0.04, 0.08) in the probability of depression, an increase of 0.53 points in ratings of poor quality of life (95%CI 0.38, 0.69), and an increase of 0.10 points in loneliness (95%CI 0.04, 0.16), corresponding respectively to a change of 26.2%, 2.4%, and 1.8%. Further, the probability of anxiety increased by almost two percentage points (95%CI 0.004, 0.027) during the second COVID-19 wave, indicating an increase of 16.6% compared with the levels of anxiety at the first COVID-19 wave. Of note, the increase in the levels of depression during COVID-19 was considerably larger than the change in the other outcomes also when considering the total number of depressive symptoms (i.e. 45% increase before COVID-19 vs COVID-19 wave 1, and 13% increase COVID-19 wave 1 vs wave 2; see SI Appendix – sTable5).

The standardised change scores of the mental health outcomes before and during COVID-19 are illustrated in Figure2. As suggested by the previous results, the sharpest deterioration before and during COVID-19 (wave 1) was found for depression (standardised change 0.26), followed by poor quality of life (standardised change 0.15) and loneliness (standardised change 0.08). Depression also showed the largest increase across the two COVID-19 waves (standardised change 0.15), followed by poor quality of life (standardised change 0.09), anxiety (standardised change 0.06), and loneliness (standardised change 0.05).

**Figure2.**
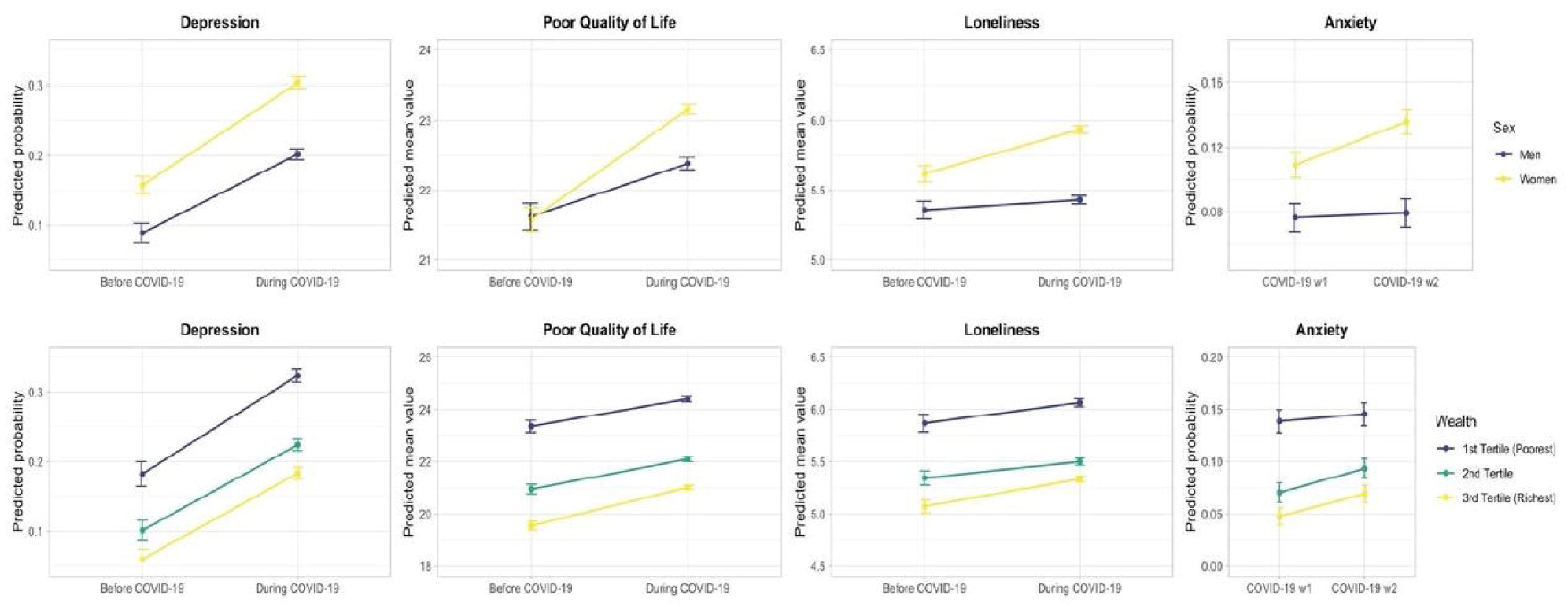
Standardised changes in mental health before and during the COVID-19 pandemic (fixed-effects models). **Note**. ELSA COVID-19 longitudinal sample (N=5,146); weighted pooled estimates from two-way fixed-effects linear models across 20 imputed datasets.

#### (4) Mental health and wellbeing impact of the COVID-19 pandemic across different sociodemographic groups

We further tested interaction effects between the COVID-19 period indicator representing the average mental health change during COVID-19 (i.e. average change across the two COVID-19 waves) and four sociodemographic characteristics – namely, sex, wealth, partnership, and age. The results (SI Appendix – sTable4) provide evidence for some heterogeneity in the mental health impact of the pandemic. Figure3 shows the predicted values of the mental health outcomes by sex and wealth, derived from the mutually adjusted interaction models. Women experienced worse changes in mental health than men across all outcomes. The increase in depression was 3 percentage points higher in women than in men (95% CI 0.004, 0.063). Average ratings of poor quality of life were 0.86 points higher in women compared with men (95%CI 0.48, 1.24). The most notable sex differences were found for loneliness (interaction effect = 0.24, 95%CI 0.11, 0.36) and anxiety (interaction effect = 0.027, 95% CI 0.003, 0.051), which increased among women but remained almost stable in men (Figure3). The increase in poor quality of life and loneliness during the pandemic was smaller for participants in the poorest wealth group compared with those in the richest wealth group (Poor Quality of Life: interaction effect = -0.53, 95%CI -0.95, -0.11; Loneliness: interaction effect = -0.15, 95%CI -0.29, 0.00). However, the levels of mental ill-health levels in the wealthiest did not reach those of people with lower wealth. It is important to note that participants with lower wealth had worse mental health than those with higher wealth both before and during COVID-19 across all outcomes considered in the analyses (Figure3); this suggests that socioeconomic inequalities in mental health have persisted during the pandemic. Further, ratings of loneliness increased significantly among participants who did not have a partner (and were living alone) (interaction effect = 0.23, 95%CI 0.07, 0.39), but remained almost stable in those who had a partner (SI Appendix – sFigure2). We did not find marked age differences in the impact of the pandemic on the mental health of older adults, as all age groups considered in the analysis (age range 52-99 years) showed a similar deterioration in mental health during COVID-19. The only exception was for depression (SI Appendix – sFigure2), which showed a smaller increase in the 75+ age group than in the youngest age group (50-59 years) (interaction effect = -0.05, 95%CI -0.09, -0.01).

**Figure3.**
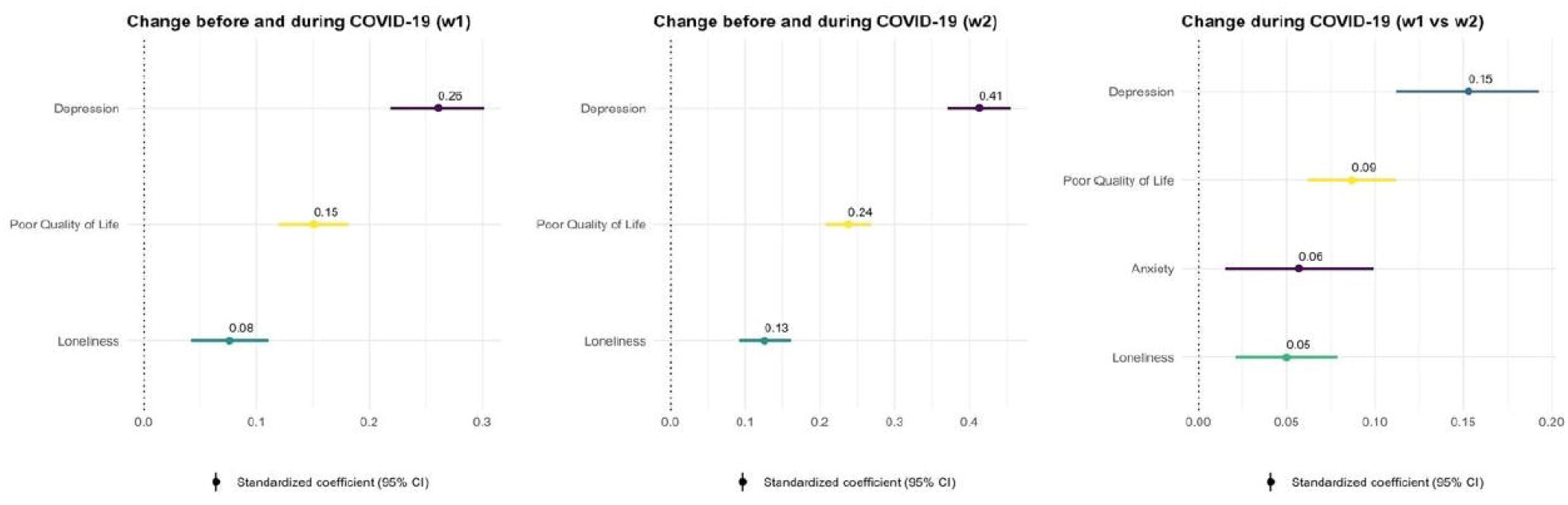
Fixed-effects models: Interaction effects between changes in mental health and sex (upper panels) and wealth (lower panels). **Note**. ELSA COVID-19 longitudinal sample (N=5,146); predicted values of the outcomes by sociodemographic characteristics, derived from mutually adjusted two-way fixed-effects linear models; weighted pooled estimates across 20 imputed datasets.

### Sensitivity analyses

First, we tested the impact of the COVID-19 pandemic and the interaction effects with sociodemographic factors on depression and anxiety using the total CESD-8 and GAD-7 scores, rather than the binary scores representing cases of depression and anxiety. The results mirrored those of the analysis with the binary scores (SI Appendix – sTable5 and sTable6). The change in the total score of depression before and during COVID-19 was smaller than the change in the binary depression score, but still considerably larger than the change observed for the total scores of quality of life and loneliness. The changes in the total and binary scores of anxiety were broadly similar (SI Appendix – sTable3 and sTable5). Second, we reran all models presented in the main imputed data analysis using the sample of participants with complete data on all variables. The pattern of changes in mental health before and during COVID-19 and interaction effects with sociodemographic factors aligned closely with the results found in the main imputed analysis (SI Appendix – sTable7 and sTable8). Lastly, we restricted the main analyses to participants who did not experience COVID-19 at either the first or second COVID-19 wave A total of 5.4% experienced COVID-19 on the basis of testing, hospitalisation, or symptoms, leaving 4,867 (94.6%) in these analyses. There were no substantial changes from the primary analyses in the magnitude and statistical significance of the associations, except for the sex interaction on anxiety which was smaller and non-significant when excluding participants who experienced COVID-19 (SI Appendix – sTable9 and sTable10).

## DISCUSSION

Using a nationally representative sample of older people living in private households in England we investigated longitudinal changes in mental health before and during the initial and later phases of the COVID-19 pandemic. Our results provided clear evidence for an overall deterioration in all mental health outcomes, which persisted throughout the course of the pandemic in 2020. Levels of depression, poor quality of life, and loneliness increased significantly during June-July and again in November-December 2020, compared with pre-pandemic levels. The largest change was observed for depression, followed by poor quality of life and loneliness. We also found a significant increase in the levels of anxiety during the pandemic. Furthermore, we showed that changes in mental health varied across distinct sociodemographic groups. Deterioration of mental health was greater in women than in men across all outcomes. Participants with less wealth had lower levels of mental health than those in the highest wealth group, before and during the pandemic. Nevertheless, people with higher wealth experienced more negative changes in quality of life and loneliness throughout the pandemic. Average ratings of loneliness increased among people who did not have a partner and were living alone, but remained relatively stable in those who had a partner. The changes in most mental health outcomes were similar across different age groups, suggesting that older people were no more resilient to the mental health impact of the pandemic than younger adults.

The novelty of this study lies in the analysis of changes in mental health outcomes among older people living in the community before and during early and later stages of the COVID-19 pandemic, and identification of vulnerable groups therein. Studies involving repeated measures have suggested that the highest levels of distress were experienced early in the pandemic, with recovery during the summer months of 2020. ^35^ We showed that mental health and wellbeing continued to worsen as lockdown continued. Recent studies on older people have not been able to analyse large nationally representative samples with pre-COVID-19 data ^5 36 37^. Therefore, our results showing lower levels of mental health during the summer and again in Autumns 2020 compared to pre-pandemic data are novel. Although it is not possible to make direct comparisons with previous studies due to differences in samples and/or follow-up periods, our result regarding vulnerable groups, i.e. women, and single/widowed/divorced, are in line with other studies ^4 8 9 36-38^. Contrary to longitudinal analyses indicating that the impact on mental health has been smaller among older than younger adults, ^9 38^ our study showed that changes in wellbeing and anxiety were similar across different age groups, but a slightly higher increase in depression was experienced in those aged 75+ than people the youngest age group (52-59 years). It is possible that this group of people continued to experience more stressors as further lockdown measures were included and did not have the time to adapt to circumstances.

The challenges to mental health of the COVID-19 pandemic take two forms: distress related to the disease itself (fear of infection, hospitalisation and death for the individual and their loved ones), and distress arising from the containment actions taken by governments (stay at home orders, social distancing, concern about finances, employment, access to services and commodities, interruption of care). Older people are at increased risk of serious consequences following infection, and may also use health and social care services to a greater extent than younger adults, while the diminished social connections with age may increase vulnerability to isolation and loneliness. Our finding that people from high socioeconomic groups experienced a steep decline in quality of life and increased loneliness has not been previously reported. A possible explanation might be that wealthier people have been more affected by social restrictions and the cessation of social and cultural activities than people in less affluent groups, which in turn might have resulted in impaired quality of life and greater loneliness. It has also been reported that older people with wealth held in risky assets have been severely hit by fluctuations in the stock markets, ^39^. Many older individuals have private pension savings that are exposed to market risk, and findings from the ELSA Covid-19 Substudy showed that 32% of people believe the value of their pension is considerably lower than pre-crisis ^39^, which in turn can affect their overall evaluation of quality of life. However, it should be noted that despite the greater increase in mental ill-health, levels of depression, loneliness and poor quality of life did not reach those of less affluent groups.

Our study has many strengths. It is based on a nationally representative sample of older individuals living in private households in England, with pre-COVID-19 data. The response rate at both assessments was very high, and the study collected information on several mental health outcomes, socio-demographic and socio-economic factors. Therefore the results are generalisable to the English population aged 50 and over. The use of appropriate statistical modelling allowed to us to control for all unobserved confounders that vary across individuals but are constant over time (e.g. genetic susceptibility), and those that are constant across individuals but change over time. A possible limitation of our study is that the first round of data collection took place as the first lockdown in England was easing. It is possible that mental health problems were much higher in April and May 2020, and we were not able to assess this. ^4^

Nevertheless, our study shows the importance of providing resources to manage or attenuate the adverse mental health impact of the pandemic on older people living in the community. Psychological reactions to pandemics are common and include maladaptive behaviours, emotional distress and withdrawal. It is known that psychological factors play an important role in adherence to public health measures, including vaccination, and in how people cope with the threat of infection and consequent losses. We believe that policies should be in place for the immediate provision of diagnosis of mental health problems and targeted psychological interventions to support older people and in particular women, non-partnered people and those from low socioeconomic groups. Access to mental health services should be improved, especially those delivered online, through smartphone technologies, and importantly those over the phone to reach older people with poorer digital resources. As the COVID-19 crisis extends beyond 2020, there is a need to sustain the mental health of older people in the population, and to plan health and social support services as face to face contact becomes more feasible. Given the strong link between depressive symptoms and the incidence and prognosis of physical health outcomes,^40^ the associations found here may promote the deterioration of health more generally among older people in the population.

Our research provides a picture of the mental health effects of the pandemic at two important time points in 2020, compared to pre-pandemic data. However, as the crisis in England evolves longer-term monitoring of the determinants of mental health and wellbeing, and the consequences that these have on physical health will be necessary.

## Data Availability

The data used in this work can be obtained free upon registration at the UK Data service https://beta.ukdataservice.ac.uk/datacatalogue/series/series?id=200011
Further information regarding the sample design and data collection methods can be found on the study website (https://www.elsa-project.ac.uk/).

https://www.ukdataservice.ac.uk/

## Footnotes

### Contributors

PZ, EI and AS contributed equally to this work. PZ, EI and AS designed the study. EI did the statistical analysis under the supervision of PZ. PZ and EI are joint first authors with equal contribution. PZ, EI and AS drafted the manuscript. PZ, EI, PD and AS provided substantial scientific input in interpreting the results and drafting the manuscript. The corresponding author attests that all listed authors meet authorship criteria and that no others meeting the criteria have been omitted. PZ, EI and AS are the guarantors.

### Funding

ESRC/UKRI; National Institute on Aging; UK Government Departments co-ordinated by NIHR. The funders of the study had no role in study design, data collection, data analysis, data interpretation, or writing of the report. All authors had full access to the data in the study and had final responsibility for the decision to submit for publication.

### Competing interests

All authors have completed the ICMJE uniform disclosure form at www.icmje.org/coi_disclosure.pdf and declare no conflict of interests.

### Ethical approval

Ethical approval for the regular ELSA study was obtained from the National Research Ethics Service. The ELSA COVID-19 Substudy has been approved by the University College London Research Ethics Committee.

### Data Sharing

The data used in this work can be obtained free upon registration at the UK Data service https://beta.ukdataservice.ac.uk/datacatalogue/series/series?id=200011

Further information regarding the sample design and data collection methods can be found on the study website (https://www.elsa-project.ac.uk/).

The lead authors affirm that the manuscript is an honest, accurate, and transparent account of the study being reported; that no important aspects of the study have been omitted; and that any discrepancies from the study as planned (and, if relevant, registered) have been explained.

Dissemination to participants and related patient and public communities: Results of this study will be disseminated to study participants and the general public via emailed newsletters, and social media channels where available.

Provenance and peer review: Not commissioned; externally peer reviewed.

## Notes

### Competing Interest Statement

The authors have declared no competing interest.

