## Supplementary materials for "The immediate and longer-term impact of the COVID-19 pandemic on the mental health and wellbeing of older adults in England"

**sTable1. Descriptive statistics of the mental health outcomes before and during COVID-19.**

| | **Imputed data, weighted** | | |
|---|---|---|---|
| | **Before COVID-19** | **During COVID-19 (w1)** | **During COVID-19 (w2)** |
| **Elevated depressive symptoms (CESD-8 ≥ 4)** | | | |
| No | 87.50% | 77.40% | 71.50% |
| Yes | 12.50% | 22.60% | 28.50% |
| **Poor Quality of Life (CASP-12)** | | | |
| Mean (SD) | 21.602 (6.283) | 22.529 (6.526) | 23.062 (6.670) |
| Range | 1.000 - 48.000 | 12.000 - 47.000 | 9.000 - 48.000 |
| **Loneliness** | | | |
| Mean (SD) | 5.496 (2.026) | 5.648 (2.065) | 5.748 (2.172) |
| Range | 1.000 - 12.000 | 2.000 - 12.000 | 3.000 - 12.000 |
| **Anxiety (Gad-7 ≥ 10)** | | | |
| No | NA | 90.60% | 89.10% |
| Yes | NA | 9.40% | 10.90% |

***Note.*** *ELSA COVID-19 longitudinal sample (N=5146); SD = standard deviation; NA = Not Available.*

**sTable2. Predicted values of the mental health outcomes before and during COVID-19 (fixed-effects model results).**

| **Outcome** | **Wave** | **Predicted values** | **Observed values** |
|---|---|---|---|
| | | **%** | **%** |
| Depression | Before COVID-19 | 12.5 | 12.5 |
| | COVID-19 w1 | 22.6 | 22.6 |
| | COVID-19 w2 | 28.5 | 28.5 |
| Anxiety | COVID-19 w1 | 9.4 | 9.4 |
| | COVID-19 w2 | 10.9 | 10.9 |
| | | **Mean (s.e.)** | **Mean(SD)** |
| Poor QoL (total score) | Before COVID-19 | 21.6 (0.06) | 21.6 (6.28) |
| | COVID-19 w1 | 22.5 (0.05) | 22.5 (6.53) |
| | COVID-19 w2 | 23.1(0.05) | 23.1 (6.67) |
| Loneliness (total score) | Before COVID-19 | 5.5 (0.02) | 5.5 (2.03) |
| | COVID-19 w1 | 5.6 (0.02) | 5.6 (2.07) |
| | COVID-19 w2 | 5.7 (0.02) | 5.7 (2.17) |

***Note**. ELSA COVID-19 longitudinal sample (N=5,146); weighted pooled estimates from two-way fixed-effects linear models across 20 imputed datasets. SE = standard error. SD = standard deviation. QoL = quality of life.*

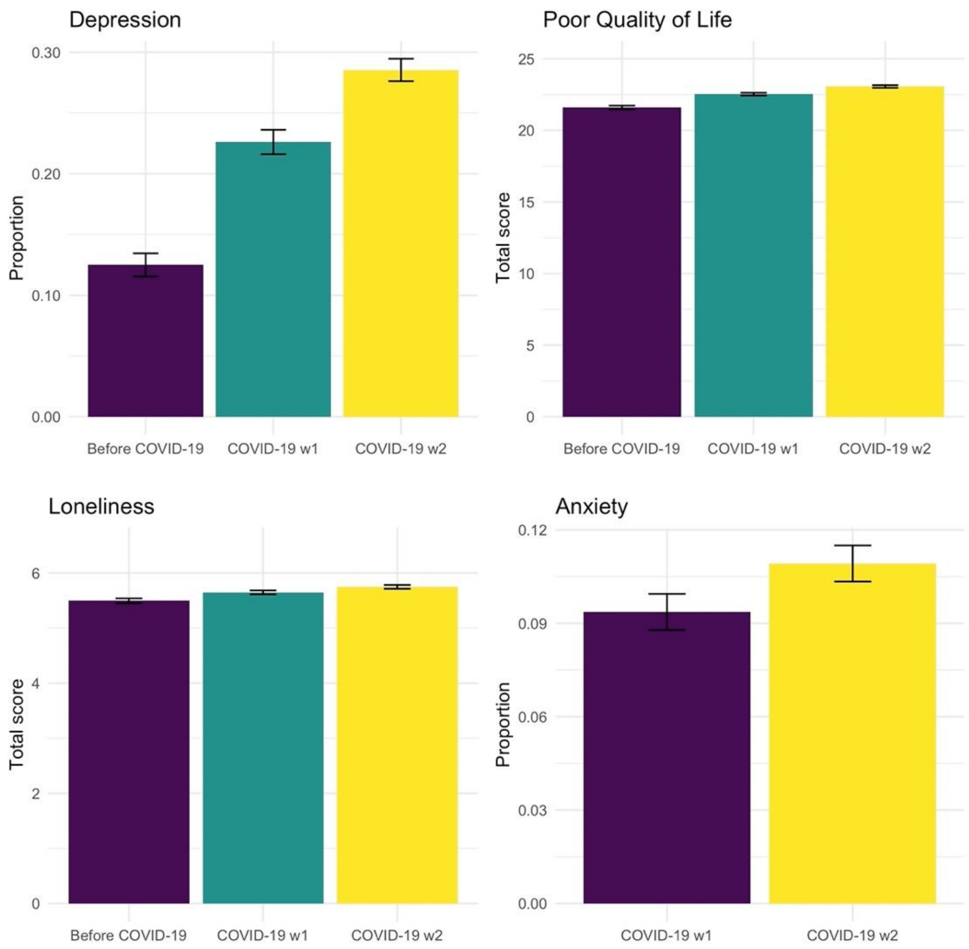


**sFigure1. Predicted values of the mental health outcomes before and during the COVID-19 pandemic.**
***Note.*** *ELSA COVID-19 longitudinal sample (N=5146); weighted pooled estimates from two-way fixed-effects linear models across 20 imputed datasets.*

| sTable3. Two-way fixed-effects models: Changes in mental health before and during the COVID-19 pandemic. | | | | | | |
|---|---|---|---|---|---|---|
| | **B** | **SE** | **p-value** | **CI (lower)** | **CI (upper)** | **% change[a]** |
| **Outcome: Depression** | | | | | | |
| Change before vs during COVID-19 (average) | 0.131 | 0.007 | **0.000** | 0.117 | 0.145 | 104.75 |
| Change before vs during COVID-19 (w1) | 0.101 | 0.008 | **0.000** | 0.085 | 0.117 | 81.02 |
| Change before vs during COVID-19 (w2) | 0.160 | 0.008 | **0.000** | 0.144 | 0.177 | 128.48 |
| Change COVID-19 w1 vs w2 | 0.059 | 0.008 | **0.000** | 0.043 | 0.075 | 26.22 |
| **Outcome: Poor Quality of Life** | | | | | | |
| Change before vs during COVID-19 (average) | 1.194 | 0.090 | **0.000** | 1.018 | 1.370 | 5.53 |
| Change before vs during COVID-19 (w1) | 0.927 | 0.099 | **0.000** | 0.732 | 1.122 | 4.29 |
| Change before vs during COVID-19 (w2) | 1.461 | 0.096 | **0.000** | 1.272 | 1.649 | 6.76 |
| Change COVID-19 w1 vs w2 | 0.534 | 0.078 | **0.000** | 0.381 | 0.686 | 2.37 |
| **Outcome: Loneliness** | | | | | | |
| Change before vs during COVID-19 (average) | 0.202 | 0.032 | **0.000** | 0.140 | 0.265 | 3.68 |
| Change before vs during COVID-19 (w1) | 0.152 | 0.035 | **0.000** | 0.083 | 0.221 | 2.77 |
| Change before vs during COVID-19 (w2) | 0.252 | 0.035 | **0.000** | 0.183 | 0.322 | 4.59 |
| Change COVID-19 w1 vs w2 | 0.100 | 0.029 | **0.001** | 0.043 | 0.157 | 1.77 |
| **Outcome: Anxiety** | | | | | | |
| Change COVID-19 w1 vs w2 | 0.016 | 0.006 | **0.008** | 0.004 | 0.027 | 16.62 |

***Note.*** ELSA COVID-19 longitudinal sample (N=5146); weighted pooled estimates across 20 imputed datasets; p-values highlighted in bold are statistically significant at the 95% confidence level. [a] Calculated as change score (i.e. slope) divided by baseline value (i.e. intercept) and multiplied by 100%.

**sTable4. Two-way fixed-effects models: Interaction effects between changes in mental health and sociodemographic characteristics.**

| | | Outcome: Depression | | | | | Outcome: Poor Quality of Life | | | | | Outcome: Loneliness | | | | | Outcome: Anxiety | | | | |
|---|---|---|---|---|---|---|---|---|---|---|---|---|---|---|---|---|---|---|---|---|---|
| **Sociodemographic characteristics** | | **B** | **SE** | **p-value** | **CI (lower)** | **CI (upper)** | **B** | **SE** | **p-value** | **CI (lower)** | **CI (upper)** | **B** | **SE** | **p-value** | **CI (lower)** | **CI (upper)** | **B** | **SE** | **p-value** | **CI (lower)** | **CI (upper)** |
| **a. Individual interactions** | | | | | | | | | | | | | | | | | | | | | |
| *Age group* | Change | 0.149 | 0.015 | **0.000** | 0.119 | 0.180 | 1.348 | 0.195 | **0.000** | 0.967 | 1.730 | 0.251 | 0.073 | **0.001** | 0.108 | 0.394 | 0.016 | 0.006 | **0.008** | 0.004 | 0.027 |
| | Change*50-59 | ref | | | | | ref | | | | | ref | | | | | ref | | | | |
| | Change*60-74 | -0.017 | 0.019 | 0.367 | -0.053 | 0.020 | -0.319 | 0.225 | 0.157 | -0.760 | 0.123 | -0.113 | 0.083 | 0.174 | -0.275 | 0.050 | -0.017 | 0.016 | 0.295 | -0.048 | 0.014 |
| | Change*75 and over | -0.047 | 0.020 | **0.021** | -0.086 | -0.007 | -0.055 | 0.261 | 0.834 | -0.566 | 0.457 | 0.006 | 0.093 | 0.948 | -0.177 | 0.189 | -0.018 | 0.017 | 0.281 | -0.051 | 0.015 |
| *Sex* | Change | 0.113 | 0.011 | **0.000** | 0.092 | 0.134 | 0.759 | 0.143 | **0.000** | 0.480 | 1.039 | 0.073 | 0.046 | 0.115 | -0.018 | 0.164 | 0.000 | 0.000 | | 0.000 | 0.000 |
| | Change*Men | ref | | | | | ref | | | | | ref | | | | | ref | | | | |
| | Change*Women | 0.034 | 0.015 | **0.022** | 0.005 | 0.063 | 0.821 | 0.191 | **0.000** | 0.447 | 1.196 | 0.245 | 0.064 | **0.000** | 0.120 | 0.369 | 0.024 | 0.012 | **0.043** | 0.001 | 0.047 |
| *Wealth (tertiles)* | Change | 0.123 | 0.011 | **0.000** | 0.102 | 0.145 | 1.465 | 0.124 | **0.000** | 1.221 | 1.708 | 0.262 | 0.044 | **0.000** | 0.176 | 0.348 | 0.000 | 0.000 | | 0.000 | 0.000 |
| | Change*1st tertile | 0.018 | 0.017 | 0.314 | -0.017 | 0.052 | -0.417 | 0.207 | **0.044** | -0.823 | -0.011 | -0.065 | 0.073 | 0.374 | -0.209 | 0.079 | -0.015 | 0.014 | 0.280 | -0.042 | 0.012 |
| | Change*2nd tertile | -0.001 | 0.015 | 0.959 | -0.031 | 0.030 | -0.309 | 0.189 | 0.103 | -0.680 | 0.062 | -0.106 | 0.067 | 0.115 | -0.237 | 0.026 | 0.001 | 0.012 | 0.907 | -0.023 | 0.026 |
| | Change*3rd tertile | ref | | | | | ref | | | | | ref | | | | | ref | | | | |
| *Partnership* | Change | 0.126 | 0.008 | **0.000** | 0.111 | 0.142 | 1.222 | 0.102 | **0.000** | 1.022 | 1.422 | 0.146 | 0.035 | **0.000** | 0.078 | 0.215 | 0.000 | 0.000 | | 0.000 | 0.000 |
| | Change*Partnered | ref | | | | | ref | | | | | ref | | | | | ref | | | | |
| | Change*Non-partnered | 0.020 | 0.019 | 0.299 | -0.017 | 0.056 | -0.116 | 0.213 | 0.586 | -0.534 | 0.302 | 0.228 | 0.080 | **0.005** | 0.070 | 0.386 | -0.022 | 0.013 | 0.097 | -0.049 | 0.004 |
| **b. Mutually adjusted interactions** | | | | | | | | | | | | | | | | | | | | | |
| | Change | 0.124 | 0.021 | **0.000** | 0.084 | 0.165 | 1.302 | 0.235 | **0.000** | 0.841 | 1.763 | 0.205 | 0.086 | **0.018** | 0.035 | 0.374 | 0.000 | 0.000 | | 0.000 | 0.000 |
| *Age group* | Change*50-59 | ref | | | | | ref | | | | | ref | | | | | ref | | | | |
| | Change*60-74 | -0.017 | 0.019 | 0.370 | -0.054 | 0.020 | -0.386 | 0.226 | 0.087 | -0.828 | 0.057 | -0.143 | 0.083 | 0.084 | -0.306 | 0.019 | -0.018 | 0.016 | 0.246 | -0.049 | 0.013 |
| | Change*75 and over | -0.053 | 0.020 | **0.007** | -0.092 | -0.014 | -0.121 | 0.275 | 0.659 | -0.660 | 0.418 | -0.072 | 0.096 | 0.455 | -0.261 | 0.117 | -0.017 | 0.017 | 0.296 | -0.050 | 0.015 |
| *Sex* | Change*Men | ref | | | | | ref | | | | | ref | | | | | ref | | | | |
| | Change*Women | 0.033 | 0.015 | **0.026** | 0.004 | 0.063 | 0.856 | 0.194 | **0.000** | 0.476 | 1.236 | 0.235 | 0.064 | **0.000** | 0.110 | 0.360 | 0.027 | 0.012 | **0.026** | 0.003 | 0.051 |
| *Wealth (tertiles)* | Change*1st tertile | 0.008 | 0.018 | 0.659 | -0.027 | 0.043 | -0.528 | 0.214 | **0.013** | -0.947 | -0.110 | -0.145 | 0.074 | **0.050** | -0.290 | 0.000 | -0.017 | 0.014 | 0.236 | -0.044 | 0.011 |
| | Change*2nd tertile | -0.002 | 0.016 | 0.906 | -0.033 | 0.029 | -0.351 | 0.189 | 0.064 | -0.722 | 0.020 | -0.137 | 0.067 | **0.043** | -0.269 | -0.004 | 0.001 | 0.012 | 0.913 | -0.023 | 0.026 |
| | Change*3rd tertile | ref | | | | | ref | | | | | ref | | | | | ref | | | | |
| *Partnership* | Change*Partnered | ref | | | | | ref | | | | | ref | | | | | ref | | | | |
| | Change*Non-partnered | 0.025 | 0.019 | 0.194 | -0.013 | 0.062 | -0.122 | 0.227 | 0.591 | -0.567 | 0.323 | 0.230 | 0.084 | **0.006** | 0.066 | 0.394 | -0.020 | 0.014 | 0.162 | -0.047 | 0.008 |

***Note.*** ELSA COVID-19 longitudinal sample (N=5146); weighted pooled estimates across 20 imputed datasets; p-values highlighted in bold are statistically significant at the 95% confidence level; interaction effects of sociodemographic characteristics with change before and/or during COVID-19 (average).

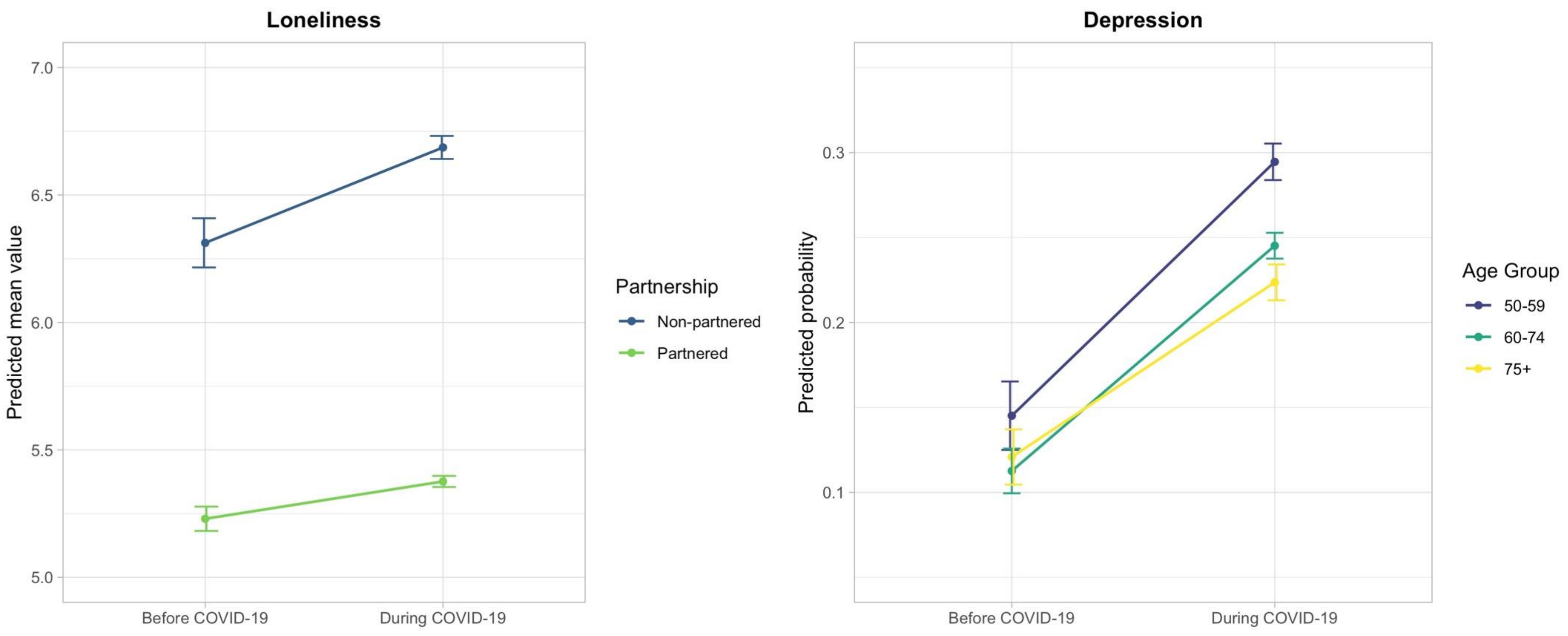


**sFigure2. Fixed-effects models: Interaction effects between changes in mental health and sociodemographic characteristics.**
***Note.*** ELSA COVID-19 longitudinal sample (N=5146); predicted values of the outcomes by sociodemographic characteristics, derived from mutually adjusted two-way fixed-effects linear models; weighted pooled estimates across 20 imputed datasets.

**sTable5. Two-way fixed-effects models: Changes in the total scores of depression and anxiety before and during the COVID-19 pandemic.**

| | B | SE | p-value | CI (lower) | CI (upper) | % change[a] |
|---|---|---|---|---|---|---|
| **Outcome: Depression** | | | | | | |
| Change before vs during COVID-19 (average) | 0.774 | 0.037 | **0.000** | 0.702 | 0.846 | 55.01 |
| Change before vs during COVID-19 (w1) | 0.638 | 0.040 | **0.000** | 0.559 | 0.717 | 45.33 |
| Change before vs during COVID-19 (w2) | 0.911 | 0.041 | **0.000** | 0.831 | 0.991 | 64.68 |
| Change COVID-19 w1 vs w2 | 0.272 | 0.035 | **0.000** | 0.204 | 0.341 | 13.31 |
| **Outcome: Anxiety** | | | | | | |
| Change COVID-19 w1 vs w2 | 0.369 | 0.070 | **0.000** | 0.230 | 0.507 | 11.22 |

***Note.*** ELSA COVID-19 longitudinal sample (N=5146); weighted pooled estimates across 20 imputed datasets; p-values highlighted in bold are statistically significant at the 95% confidence level. [a] Calculated as change score (i.e. slope) divided by baseline value (i.e. intercept) and multiplied by 100%.

| **sTable6. Two-way fixed-effects models: Interaction effects between changes in the total scores of depression and anxiety and sociodemographic characteristics.** | | | | | | | | | | | |
|---|---|---|---|---|---|---|---|---|---|---|---|
| | | **Outcome: Depression** | | | | | **Outcome: Anxiety** | | | | |
| **Sociodemographic characteristics** | | **B** | **SE** | **p-value** | **CI (lower)** | **CI (upper)** | **B** | **SE** | **p-value** | **CI (lower)** | **CI (upper)** |
| **a. Individual interactions** | | | | | | | | | | | |
| *Age group* | Change | 0.919 | 0.078 | **0.000** | 0.767 | 1.071 | 0.510 | 0.160 | **0.001** | 0.197 | 0.823 |
| | Change*50-59 | ref | | | | | ref | | | | |
| | Change*60-74 | -0.132 | 0.094 | 0.160 | -0.315 | 0.052 | -0.183 | 0.185 | 0.324 | -0.546 | 0.181 |
| | Change*75 and over | -0.363 | 0.098 | **0.000** | -0.555 | -0.172 | -0.254 | 0.192 | 0.185 | -0.630 | 0.122 |
| *Sex* | Change | 0.678 | 0.056 | **0.000** | 0.568 | 0.788 | 0.263 | 0.108 | **0.015** | 0.051 | 0.475 |
| | Change*Men | ref | | | | | ref | | | | |
| | Change*Women | 0.183 | 0.074 | **0.014** | 0.037 | 0.329 | 0.200 | 0.142 | 0.159 | -0.078 | 0.479 |
| *Wealth (tertiles)* | Change | 0.722 | 0.055 | **0.000** | 0.615 | 0.829 | 0.398 | 0.106 | **0.000** | 0.190 | 0.607 |
| | Change*1st tertile | 0.094 | 0.088 | 0.285 | -0.078 | 0.266 | -0.054 | 0.168 | 0.747 | -0.383 | 0.275 |
| | Change*2nd tertile | 0.041 | 0.079 | 0.605 | -0.114 | 0.195 | -0.022 | 0.152 | 0.887 | -0.320 | 0.277 |
| | Change*3rd tertile | ref | | | | | ref | | | | |
| *Partnership* | Change | 0.739 | 0.040 | **0.000** | 0.661 | 0.818 | 0.366 | 0.080 | **0.000** | 0.209 | 0.523 |
| | Change*Partnered | ref | | | | | ref | | | | |
| | Change*Non-partnered | 0.143 | 0.094 | 0.130 | -0.042 | 0.328 | 0.009 | 0.168 | 0.957 | -0.320 | 0.339 |
| **b. Mutually adjusted interactions** | | | | | | | | | | | |
| | Change | 0.771 | 0.100 | **0.000** | 0.575 | 0.968 | 0.461 | 0.199 | **0.020** | 0.071 | 0.851 |
| *Age group* | Change*50-59 | ref | | | | | ref | | | | |
| | Change*60-74 | -0.140 | 0.094 | 0.139 | -0.325 | 0.045 | -0.202 | 0.187 | 0.280 | -0.569 | 0.165 |
| | Change*75 and over | -0.422 | 0.099 | **0.000** | -0.615 | -0.229 | -0.292 | 0.193 | 0.131 | -0.671 | 0.087 |
| *Sex* | Change*Men | ref | | | | | ref | | | | |
| | Change*Women | 0.183 | 0.076 | **0.016** | 0.035 | 0.332 | 0.212 | 0.146 | 0.146 | -0.074 | 0.498 |
| *Wealth (tertiles)* | Change*1st tertile | 0.023 | 0.089 | 0.799 | -0.152 | 0.198 | -0.112 | 0.174 | 0.518 | -0.453 | 0.228 |
| | Change*2nd tertile | 0.033 | 0.079 | 0.675 | -0.122 | 0.189 | -0.033 | 0.154 | 0.828 | -0.335 | 0.268 |
| | Change*3rd tertile | ref | | | | | ref | | | | |
| *Partnership* | Change*Partnered | ref | | | | | ref | | | | |
| | Change*Non-partnered | 0.200 | 0.098 | **0.041** | 0.008 | 0.391 | 0.053 | 0.175 | 0.762 | -0.290 | 0.396 |
| ***Note.*** ELSA COVID-19 longitudinal sample (N=5146); weighted pooled estimates across 20 imputed datasets; p-values highlighted in bold are statistically significant at the 95% confidence level. | | | | | | | | | | | |

| **sTable7. Two-way fixed-effects models: Changes in mental health before and during the COVID-19 pandemic - Complete data analysis.** | | | | | |
|---|---|---|---|---|---|
| | **B** | **SE** | **p-value** | **CI (lower)** | **CI (upper)** |
| **Outcome: Depression (N=5047)** | | | | | |
| Change before vs during COVID-19 (average) | 0.130 | 0.007 | **0.000** | 0.116 | 0.144 |
| Change before vs during COVID-19 (w1) | 0.100 | 0.007 | **0.000** | 0.085 | 0.115 |
| Change before vs during COVID-19 (w2) | 0.160 | 0.008 | **0.000** | 0.144 | 0.177 |
| Change COVID-19 w1 vs w2 | 0.061 | 0.007 | **0.000** | 0.046 | 0.075 |
| **Outcome: Poor Quality of Life (N=5047)** | | | | | |
| Change before vs during COVID-19 (average) | 1.194 | 0.083 | **0.000** | 1.031 | 1.357 |
| Change before vs during COVID-19 (w1) | 0.927 | 0.093 | **0.000** | 0.744 | 1.110 |
| Change before vs during COVID-19 (w2) | 1.461 | 0.090 | **0.000** | 1.284 | 1.638 |
| Change COVID-19 w1 vs w2 | 0.534 | 0.078 | **0.000** | 0.381 | 0.686 |
| **Outcome: Loneliness (N=5047)** | | | | | |
| Change before vs during COVID-19 (average) | 0.202 | 0.029 | **0.000** | 0.145 | 0.260 |
| Change before vs during COVID-19 (w1) | 0.152 | 0.033 | **0.000** | 0.088 | 0.217 |
| Change before vs during COVID-19 (w2) | 0.252 | 0.033 | **0.000** | 0.188 | 0.317 |
| Change COVID-19 w1 vs w2 | 0.100 | 0.029 | **0.001** | 0.043 | 0.157 |
| **Outcome: Anxiety (N=5106)** | | | | | |
| Change COVID-19 w1 vs w2 | 0.016 | 0.006 | **0.008** | 0.004 | 0.027 |
| ***Note.*** Complete data analysis; weighted estimates; p-values highlighted in bold are statistically significant at the 95% confidence level. | | | | | |

**sTable8. Two-way fixed-effects models: Interaction effects between changes in mental health and sociodemographic characteristics - Complete data analysis.**

| | | **Outcome: Depression (N=5047)** | | | | | **Outcome: Poor Quality of Life (N=5047)** | | | | | **Outcome: Loneliness (N=5047)** | | | | | **Outcome: Anxiety (N=5106)** | | | | |
|---|---|---|---|---|---|---|---|---|---|---|---|---|---|---|---|---|---|---|---|---|---|
| **Sociodemographic characteristics** | | **B** | **SE** | **p-value** | **CI (lower)** | **CI (upper)** | **B** | **SE** | **p-value** | **CI (lower)** | **CI (upper)** | **B** | **SE** | **p-value** | **CI (lower)** | **CI (upper)** | **B** | **SE** | **p-value** | **CI (lower)** | **CI (upper)** |
| **a. Individual interactions** | | | | | | | | | | | | | | | | | | | | | |
| *Age group* | Change | 0.149 | 0.015 | **0.000** | 0.120 | 0.178 | 1.350 | 0.171 | **0.000** | 1.016 | 1.685 | 0.251 | 0.065 | **0.000** | 0.123 | 0.379 | 0.027 | 0.014 | 0.053 | 0.000 | 0.055 |
| | Change*50-59 | ref | | | | | ref | | | | | ref | | | | | ref | | | | |
| | Change*60-74 | -0.017 | 0.018 | 0.326 | -0.052 | 0.017 | -0.322 | 0.202 | 0.111 | -0.718 | 0.074 | -0.113 | 0.075 | 0.132 | -0.260 | 0.034 | -0.017 | 0.016 | 0.293 | -0.048 | 0.014 |
| | Change*75 and over | -0.047 | 0.019 | **0.014** | -0.084 | -0.010 | -0.057 | 0.242 | 0.815 | -0.531 | 0.418 | 0.006 | 0.085 | 0.942 | -0.160 | 0.172 | -0.018 | 0.017 | 0.279 | -0.051 | 0.015 |
| *Sex* | Change | 0.112 | 0.010 | **0.000** | 0.092 | 0.133 | 0.759 | 0.132 | **0.000** | 0.500 | 1.019 | 0.073 | 0.042 | 0.086 | -0.010 | 0.155 | 0.003 | 0.009 | 0.750 | -0.015 | 0.020 |
| | Change*Men | ref | | | | | ref | | | | | ref | | | | | ref | | | | |
| | Change*Women | 0.033 | 0.014 | **0.018** | 0.006 | 0.061 | 0.821 | 0.168 | **0.000** | 0.491 | 1.151 | 0.245 | 0.059 | **0.000** | 0.130 | 0.360 | 0.024 | 0.012 | **0.042** | 0.001 | 0.047 |
| *Wealth (tertiles)* | Change | 0.123 | 0.010 | **0.000** | 0.103 | 0.143 | 1.466 | 0.115 | **0.000** | 1.242 | 1.691 | 0.262 | 0.039 | **0.000** | 0.185 | 0.338 | 0.022 | 0.008 | **0.008** | 0.006 | 0.037 |
| | Change*1st tertile | 0.018 | 0.017 | 0.287 | -0.015 | 0.050 | -0.421 | 0.191 | **0.028** | -0.795 | -0.046 | -0.066 | 0.067 | 0.325 | -0.197 | 0.065 | -0.015 | 0.014 | 0.277 | -0.041 | 0.012 |
| | Change*2nd tertile | -0.001 | 0.015 | 0.955 | -0.029 | 0.028 | -0.310 | 0.175 | 0.076 | -0.652 | 0.032 | -0.106 | 0.061 | 0.083 | -0.225 | 0.014 | 0.001 | 0.012 | 0.905 | -0.022 | 0.025 |
| | Change*3rd tertile | ref | | | | | ref | | | | | ref | | | | | ref | | | | |
| *Partnership* | Change | 0.125 | 0.008 | **0.000** | 0.111 | 0.140 | 1.222 | 0.094 | **0.000** | 1.039 | 1.406 | 0.146 | 0.032 | **0.000** | 0.084 | 0.208 | 0.021 | 0.007 | **0.002** | 0.008 | 0.035 |
| | Change*Partnered | ref | | | | | ref | | | | | ref | | | | | ref | | | | |
| | Change*Non-partnered | 0.020 | 0.018 | 0.286 | -0.016 | 0.055 | -0.116 | 0.202 | 0.567 | -0.512 | 0.281 | 0.229 | 0.076 | **0.002** | 0.081 | 0.377 | -0.022 | 0.013 | 0.097 | -0.049 | 0.004 |
| **b. Mutually adjusted interactions** | | | | | | | | | | | | | | | | | | | | | |
| | Change | 0.124 | 0.019 | **0.000** | 0.088 | 0.161 | 1.306 | 0.214 | **0.000** | 0.886 | 1.726 | 0.204 | 0.074 | **0.006** | 0.060 | 0.349 | 0.025 | 0.016 | 0.126 | -0.007 | 0.057 |
| *Age group* | Change*50-59 | ref | | | | | ref | | | | | ref | | | | | ref | | | | |
| | Change*60-74 | -0.017 | 0.018 | 0.327 | -0.052 | 0.017 | -0.390 | 0.205 | 0.057 | -0.792 | 0.012 | -0.144 | 0.074 | 0.054 | -0.290 | 0.003 | -0.018 | 0.016 | 0.244 | -0.049 | 0.013 |
| | Change*75 and over | -0.054 | 0.019 | **0.004** | -0.091 | -0.017 | -0.124 | 0.259 | 0.632 | -0.631 | 0.383 | -0.072 | 0.088 | 0.414 | -0.245 | 0.101 | -0.018 | 0.017 | 0.293 | -0.050 | 0.015 |
| *Sex* | Change*Men | ref | | | | | ref | | | | | ref | | | | | ref | | | | |
| | Change*Women | 0.033 | 0.014 | **0.021** | 0.005 | 0.061 | 0.856 | 0.171 | **0.000** | 0.520 | 1.192 | 0.236 | 0.059 | **0.000** | 0.120 | 0.351 | 0.027 | 0.012 | **0.025** | 0.003 | 0.051 |
| *Wealth (tertiles)* | Change*1st tertile | 0.008 | 0.017 | 0.640 | -0.025 | 0.041 | -0.532 | 0.199 | **0.007** | -0.922 | -0.143 | -0.145 | 0.067 | **0.030** | -0.277 | -0.014 | -0.017 | 0.014 | 0.232 | -0.044 | 0.011 |
| | Change*2nd tertile | -0.002 | 0.015 | 0.898 | -0.031 | 0.027 | -0.352 | 0.174 | **0.043** | -0.694 | -0.011 | -0.137 | 0.061 | **0.025** | -0.256 | -0.017 | 0.001 | 0.012 | 0.911 | -0.023 | 0.025 |
| | Change*3rd tertile | ref | | | | | ref | | | | | ref | | | | | ref | | | | |
| *Partnership* | Change*Partnered | ref | | | | | ref | | | | | ref | | | | | ref | | | | |
| | Change*Non-partnered | 0.025 | 0.019 | 0.183 | -0.012 | 0.061 | -0.121 | 0.215 | 0.574 | -0.542 | 0.301 | 0.231 | 0.079 | **0.003** | 0.076 | 0.386 | -0.020 | 0.014 | 0.161 | -0.047 | 0.008 |

***Note.*** Complete data analysis; weighted estimates; p-values highlighted in bold are statistically significant at the 95% confidence level. Interaction effects of sociodemographic characteristics with change before and during COVID-19 (average).

**sTable9. Two-way fixed-effects models: Changes in mental health before and during the COVID-19 pandemic – Models restricted to participants who did not experience COVID-19.**

| | **B** | **SE** | **p-value** | **CI (lower)** | **CI (upper)** |
|---|---|---|---|---|---|
| **Outcome: Depression** | | | | | |
| Change before vs during COVID-19 (average) | 0.129 | 0.007 | **0.000** | 0.114 | 0.143 |
| Change before vs during COVID-19 (w1) | 0.100 | 0.008 | **0.000** | 0.084 | 0.117 |
| Change before vs during COVID-19 (w2) | 0.157 | 0.009 | **0.000** | 0.140 | 0.174 |
| Change COVID-19 w1 vs w2 | 0.057 | 0.008 | **0.000** | 0.041 | 0.073 |
| **Outcome: Poor Quality of Life** | | | | | |
| Change before vs during COVID-19 (average) | 1.151 | 0.088 | **0.000** | 0.980 | 1.323 |
| Change before vs during COVID-19 (w1) | 0.885 | 0.098 | **0.000** | 0.693 | 1.078 |
| Change before vs during COVID-19 (w2) | 1.417 | 0.094 | **0.000** | 1.233 | 1.601 |
| Change COVID-19 w1 vs w2 | 0.532 | 0.078 | **0.000** | 0.378 | 0.685 |
| **Outcome: Loneliness** | | | | | |
| Change before vs during COVID-19 (average) | 0.170 | 0.032 | **0.000** | 0.107 | 0.233 |
| Change before vs during COVID-19 (w1) | 0.115 | 0.035 | **0.001** | 0.046 | 0.185 |
| Change before vs during COVID-19 (w2) | 0.225 | 0.035 | **0.000** | 0.156 | 0.294 |
| Change COVID-19 w1 vs w2 | 0.110 | 0.029 | **0.000** | 0.052 | 0.167 |
| **Outcome: Anxiety** | | | | | |
| Change COVID-19 w1 vs w2 | 0.015 | 0.006 | **0.016** | 0.003 | 0.026 |

***Note.*** ELSA COVID-19 longitudinal sample (N=4867); weighted pooled estimates across 20 imputed datasets; p-values highlighted in bold are statistically significant at the 95% confidence level.

**sTable10. Two-way fixed-effects models: Interaction effects between changes in mental health and sociodemographic characteristics – Models restricted to participants who did not experience COVID-19.**

| | | **Outcome: Depression** | | | | | **Outcome: Poor Quality of Life** | | | | | **Outcome: Loneliness** | | | | | **Outcome: Anxiety** | | | | |
|---|---|---|---|---|---|---|---|---|---|---|---|---|---|---|---|---|---|---|---|---|---|
| **Sociodemographic characteristics** | | **B** | **SE** | **p-value** | **CI (lower)** | **CI (upper)** | **B** | **SE** | **p-value** | **CI (lower)** | **CI (upper)** | **B** | **SE** | **p-value** | **CI (lower)** | **CI (upper)** | **B** | **SE** | **p-value** | **CI (lower)** | **CI (upper)** |
| **a. Individual interactions** | | | | | | | | | | | | | | | | | | | | | |
| *Age group* | Change | 0.147 | 0.016 | **0.000** | 0.115 | 0.179 | 1.347 | 0.197 | **0.000** | 0.961 | 1.733 | 0.177 | 0.075 | **0.018** | 0.030 | 0.324 | 0.024 | 0.015 | 0.113 | -0.006 | 0.053 |
| | Change*50-59 | ref | | | | | ref | | | | | ref | | | | | ref | | | | |
| | Change*60-74 | -0.018 | 0.019 | 0.352 | -0.055 | 0.020 | -0.365 | 0.228 | 0.109 | -0.811 | 0.082 | -0.047 | 0.084 | 0.577 | -0.213 | 0.118 | -0.011 | 0.017 | 0.488 | -0.044 | 0.021 |
| | Change*75 and over | -0.043 | 0.020 | **0.035** | -0.083 | -0.003 | -0.130 | 0.251 | 0.604 | -0.622 | 0.362 | 0.058 | 0.093 | 0.532 | -0.125 | 0.242 | -0.016 | 0.018 | 0.351 | -0.051 | 0.018 |
| *Sex* | Change | 0.112 | 0.011 | **0.000** | 0.091 | 0.133 | 0.651 | 0.138 | **0.000** | 0.382 | 0.921 | 0.048 | 0.047 | 0.306 | -0.044 | 0.140 | 0.006 | 0.009 | 0.537 | -0.012 | 0.024 |
| | Change*Men | ref | | | | | ref | | | | | ref | | | | | ref | | | | |
| | Change*Women | 0.031 | 0.015 | **0.035** | 0.002 | 0.061 | 0.945 | 0.178 | **0.000** | 0.596 | 1.293 | 0.230 | 0.064 | **0.000** | 0.105 | 0.356 | 0.017 | 0.012 | 0.167 | -0.007 | 0.041 |
| *Wealth (tertiles)* | Change | 0.122 | 0.011 | **0.000** | 0.100 | 0.144 | 1.459 | 0.130 | **0.000** | 1.204 | 1.713 | 0.255 | 0.045 | **0.000** | 0.166 | 0.343 | 0.022 | 0.008 | **0.008** | 0.006 | 0.038 |
| | Change*1st tertile | 0.012 | 0.018 | 0.496 | -0.023 | 0.047 | -0.547 | 0.208 | **0.009** | -0.954 | -0.139 | -0.131 | 0.075 | 0.082 | -0.279 | 0.017 | -0.018 | 0.014 | 0.213 | -0.046 | 0.010 |
| | Change*2nd tertile | 0.004 | 0.016 | 0.783 | -0.027 | 0.036 | -0.261 | 0.195 | 0.179 | -0.643 | 0.120 | -0.098 | 0.067 | 0.139 | -0.229 | 0.032 | 0.002 | 0.012 | 0.899 | -0.022 | 0.025 |
| | Change*3rd tertile | ref | | | | | ref | | | | | ref | | | | | ref | | | | |
| *Partnership* | Change | 0.125 | 0.008 | **0.000** | 0.109 | 0.141 | 1.174 | 0.099 | **0.000** | 0.980 | 1.367 | 0.128 | 0.037 | **0.000** | 0.057 | 0.200 | 0.019 | 0.007 | **0.008** | 0.005 | 0.033 |
| | Change*Partnered | ref | | | | | ref | | | | | ref | | | | | ref | | | | |
| | Change*Non-partnered | 0.015 | 0.019 | 0.410 | -0.021 | 0.052 | -0.090 | 0.211 | 0.671 | -0.502 | 0.323 | 0.169 | 0.077 | **0.028** | 0.018 | 0.319 | -0.017 | 0.014 | 0.199 | -0.044 | 0.009 |
| **b. Mutually adjusted interactions** | | | | | | | | | | | | | | | | | | | | | |
| | Change | 0.125 | 0.021 | **0.000** | 0.084 | 0.166 | 1.290 | 0.254 | **0.000** | 0.792 | 1.787 | 0.164 | 0.090 | 0.068 | -0.012 | 0.341 | 0.025 | 0.017 | 0.145 | -0.009 | 0.059 |
| *Age group* | Change*50-59 | ref | | | | | ref | | | | | ref | | | | | ref | | | | |
| | Change*60-74 | -0.019 | 0.019 | 0.331 | -0.057 | 0.019 | -0.461 | 0.228 | **0.043** | -0.909 | -0.014 | -0.082 | 0.084 | 0.326 | -0.246 | 0.082 | -0.014 | 0.016 | 0.408 | -0.046 | 0.019 |
| | Change*75 and over | -0.050 | 0.020 | **0.013** | -0.090 | -0.010 | -0.244 | 0.251 | 0.332 | -0.736 | 0.249 | -0.012 | 0.093 | 0.895 | -0.195 | 0.170 | -0.017 | 0.017 | 0.328 | -0.051 | 0.017 |
| *Sex* | Change*Men | ref | | | | | ref | | | | | ref | | | | | ref | | | | |
| | Change*Women | 0.032 | 0.015 | **0.036** | 0.002 | 0.061 | 0.996 | 0.179 | **0.000** | 0.646 | 1.347 | 0.228 | 0.064 | **0.000** | 0.102 | 0.354 | 0.020 | 0.012 | 0.110 | -0.005 | 0.044 |
| *Wealth (tertiles)* | Change*1st tertile | 0.003 | 0.019 | 0.858 | -0.033 | 0.040 | -0.687 | 0.212 | **0.001** | -1.102 | -0.272 | -0.191 | 0.077 | **0.013** | -0.341 | -0.041 | -0.019 | 0.015 | 0.190 | -0.048 | 0.010 |
| | Change*2nd tertile | 0.003 | 0.016 | 0.836 | -0.028 | 0.035 | -0.318 | 0.195 | 0.104 | -0.700 | 0.065 | -0.123 | 0.067 | 0.066 | -0.255 | 0.008 | 0.001 | 0.012 | 0.904 | -0.023 | 0.026 |
| | Change*3rd tertile | ref | | | | | ref | | | | | ref | | | | | ref | | | | |
| *Partnership* | Change*Partnered | ref | | | | | ref | | | | | ref | | | | | ref | | | | |
| | Change*Non-partnered | 0.021 | 0.019 | 0.273 | -0.016 | 0.058 | -0.066 | 0.216 | 0.758 | -0.490 | 0.357 | 0.171 | 0.080 | **0.032** | 0.014 | 0.327 | -0.013 | 0.014 | 0.342 | -0.041 | 0.014 |

***Note.*** ELSA COVID-19 longitudinal sample (N=4867); weighted pooled estimates across 20 imputed datasets; p-values highlighted in bold are statistically significant at the 95% confidence level; interaction effects of sociodemographic characteristics with change before and during COVID-19 (average).
